# A Clinically Translatable Immune-based Classification of HPV-associated Head and Neck Cancer with Implications for Biomarker-Driven Treatment Deintensification and Immunotherapy

**DOI:** 10.1101/2022.02.26.22271531

**Authors:** Peter YF. Zeng, Matthew J. Cecchini, John W. Barrett, Matthew Shammas-Toma, Loris De Cecco, Mara S. Serafini, Stefano Cavalieri, Lisa Licitra, Frank Hoebers, Ruud H. Brakenhoff, C René Leemans, Kathrin Scheckenbach, Tito Poli, Xiaowei Wang, Xinyi Liu, Francisco Laxague, Eitan Prisman, Catherine Poh, Pinaki Bose, Joseph C. Dort, Mushfiq H. Shaikh, Sarah EB. Ryan, Allie Dawson, Mohammed I. Khan, Christopher J. Howlett, William Stecho, Paul Plantinga, Sabrina Daniela da Silva, Michael Hier, Halema Khan, Danielle MacNeil, Adrian Mendez, John Yoo, Kevin Fung, Pencilla Lang, Eric Winquist, David A. Palma, Hedyeh Ziai, Antonio L. Amelio, Shawn S-C. Li, Paul C. Boutros, Joe S. Mymryk, Anthony C. Nichols

## Abstract

**Purpose:** Human papillomavirus-associated (HPV^+^) head and neck squamous cell carcinoma (HNSCC) is the fastest rising cancer in North America. There is significant interest in treatment de-escalation for these patients given the generally favourable prognosis. However, 15-30% of patients recur after primary treatment, reflecting a need for improved risk-stratification tools. We sought to develop a molecular test to predict the survival of patients with newly diagnosed HPV^+^ HNSCC.

**Methods:** We created a prognostic score (UWO3) that was successfully validated in six independent cohorts comprising 906 patients, including blinded retrospective and prospective external validations. Transcriptomic data from two aggressive radiation de-escalation cohorts were used to assess the ability of UWO3 to identify patients who recur. Multivariate Cox models were used to assess the associations between the UWO3 immune class and outcomes.

**Results:** A three-gene immune score classified patients into three immune classes (immune rich, mixed, or immune desert) and was strongly associated with disease-free survival in six datasets, including large retrospective and prospective datasets. Pooled analysis demonstrated that the immune rich group had superior disease-free survival at 5 years to the immune desert (HR= 9.0, 95% CI 3.2–25.5, *P*=3.6×10^−5^) and mixed (HR=6.4, 95%CI 2.2–18.7, *P*=0.006) groups after adjusting for age, sex, smoking status, and AJCC8 clinical stage. Finally, UWO3 was able to identify patients from two treatment de-escalation cohorts who remain disease-free after aggressive de-escalation to 30 Gy radiation.

**Conclusions:** The UWO3 immune score could enable biomarker-driven clinical decision-making for patients with HPV^+^ HNSCC based on robust outcome prediction across six independent cohorts. The superior survival of immune rich patients supports de-intensification strategies, while the inferior outcomes of the immune desert patients suggest the potential for intensification and/or immunotherapy. Prospective de-escalation and intensification clinical trials are currently being planned.

## Introduction

The incidence of human papillomavirus-associated (HPV^+^) head and neck squamous cell carcinoma (HNSCC) is increasing worldwide^1^. HPV^+^ HNSCC is biologically and clinically distinct from non-HPV driven (HPV^-^) HNSCC, which is typically associated with tobacco and alcohol consumption^2-4^. Although HPV^+^ HNSCC patients are usually younger and exhibit markedly improved outcomes compared to HPV^-^ HNSCC patients^3^, current treatment guidelines from both the American Society of Clinical Oncology (ASCO) and the National Comprehensive Cancer Network recommend identical treatment regimens of high-dose cisplatin and 70Gy radiation (CRT) independent of HPV status^5,6^. The rising incidence of HPV^+^ HNSCC is leading to an increasing number of young survivors, making post-treatment quality of life and treatment-related morbidities a major concern. There is significant interest in treatment de-intensification for HPV^+^ HNSCC patients to reduce morbidity rates while maintaining the outstanding cure rates^7-9^.

Treatment de-intensification efforts have been complicated by the ∼15-30% recurrence or metastasis rate of HPV^+^ HNSCC patients treated with the current standard of care therapy^10^. Early efforts at de-intensification demonstrated that modification of current standard of care CRT can result in harm for patients. Both the De-ESCALaTE HPV and RTOG1016 phase III randomized trials found that substitution of cisplatin for cetuximab led to inferior survival outcomes^11,12^. Reflecting these unsuccessful attempts that led to poorer outcomes for patients on the experimental arm, recent treatment de-intensification guidelines from ASCO^13^ and the Head and Neck Cancer International Group^14^ have called for de-intensification to be only be attempted in the context of a clinical trial for patients with favorable risk profiles. Thus, the ideal treatment de-intensification method and patient population remain highly controversial^5,13-18^.

Molecular biomarkers reflecting the biology of the tumour may better risk-identify patients who are ideal candidates for treatment de-intensification. HPV^+^ HNSCC treatment failure has been linked to TP53 mutations^19^, tumour hypoxia^20^, keratinocyte differentiation^21^, chromosome 3p arm loss^22^, and HPV-related transcriptional programs^23^. These findings have not yet been thoroughly validated, require complex assays only available at select institutions, or generally exhibit modest effect-sizes that explain only a fraction of HPV^+^ treatment failures. There remains an urgent need for improved risk-stratification to guide therapeutic decision-making in balancing treatment toxicity and therapeutic efficacy.

We created a clinically translatable immune classification tool strongly associated with survival outcomes in HPV^+^ HNSCC based upon the abundance of three transcripts. We validated it in six HPV^+^ HNSCC cohorts comprising 906 patients, including two blinded cohorts and a tissue microarray cohort using immunohistochemistry. Finally, we show this immune classification can identify patients who respond to aggressive treatment de-escalation. Taken together, our results may enable biomarker-guided personalized treatment de-intensification and intensification in HPV^+^ HNSCC low and high-risk groups.

## Results

### Development and Validation of the UWO3 Immune Classification

To reveal transcriptomic features predictive of treatment response, we performed RNA-seq on 43 HPV^+^ HNSCC tumours, 16 of which experienced local, regional, or distant recurrence. As specific tumour microenvironment alterations are associated with recurrence in HPV^+^ HNSCC^24-27^, we characterized the tumour microenvironment (TME) by the abundance of distinct cell populations ^28^. Through unsupervised clustering of these estimated immune cell abundances (see **Methods**), we classified samples into three categories: “immune rich”, “immune desert”, and “mixed” (**Supplementary Figure S1A**). These three TME subtypes exhibited distinct patterns of overall (OS; *P =* 0.003, log rank test; **Supplementary Figure S1B**) and disease-free survival (DFS; *P* < 10^−3^, log rank test; **Supplementary Figure S1C**). To facilitate translation of these results into the clinic, we developed a minimal classifier based on using the Least Absolute Shrinkage and Selection Operator (LASSO; see **Extended Methods, Supplementary Figure S2**) to stratify patients into one of three immune classes. The resulting classifier, which we call <u>U</u>niversity of <u>W</u>estern <u>O</u>ntario 3 (UWO3), is based on the abundance of three transcripts (*CD3E, IRF4*, and *ZAP70*) and assigns immune classes strongly associated with DFS in our discovery cohort, as expected (*P* < 10^−3^, log rank test, **Supplementary Figure S3A**).

We next tested five independent cohorts to validate the association between UWO3 immune class and survival outcomes in HPV^+^ HNSCC. We first used the public The Cancer Genome Atlas (TCGA)^29^ (n = 71) and Johns Hopkins University (JHU; n = 47) HPV^+^ HNSCC RNA-seq cohorts^30^. In both, the immune rich patients had improved DFS and OS (TCGA: *P* = 0.01, JHU: *P* = 0.02, log rank test, **Figure 1A,C, Supplementary Figure S3B, C**).

**Figure 1.**
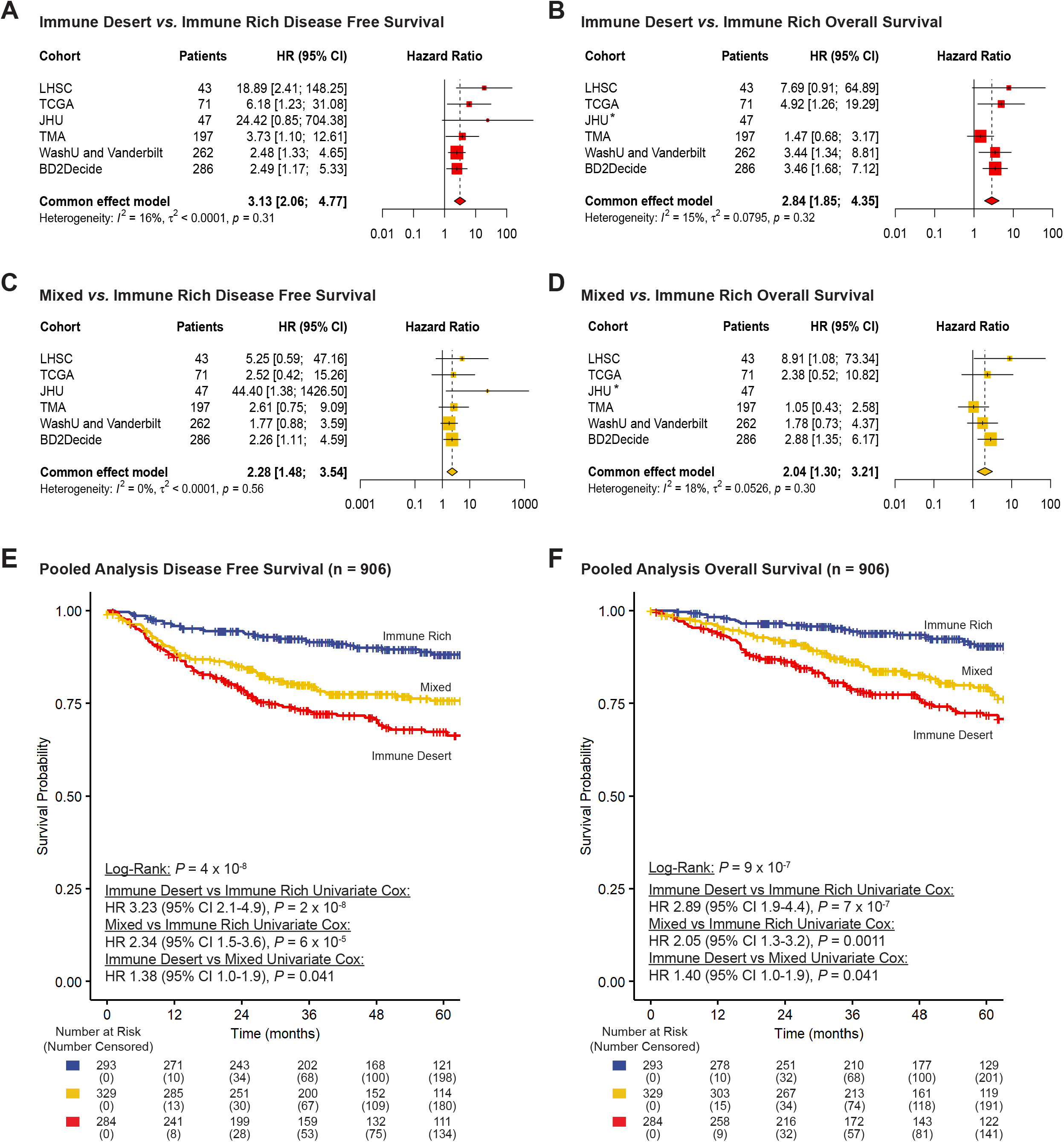
UWO3 immune group is a strong predictor of survival outcomes across six independent cohorts. Patients from the immune desert and mixed group show inferior disease free survival and overall survival compared to the immune rich patients (**A-D**). Hazard ratio (HR) from univariate Cox model and were combined using Mantel-Haenszel fixed-effect model. Heterogeneity between studies was analyzed with *χ*2 and *I*^2^ statistics. * Hazard ratio for overall survival in the JHU cohort excluded from the analysis due to only one event in the cohort. Pooled Kaplan-Meier analyses of disease-free survival (**E**) and overall survival (**F**) of HPV+ HNSCC patients show that UWO3 immune groups are associated with distinct survival outcomes. *P* values from two-sided log rank tests and Cox proportional regression model.

Next, we employed clinically-validated antibodies for the proteins corresponding to the transcripts within the UWO3 score (CD3E, ZAP70, and IRF4) that are used routinely in clinical pathology labs for hematologic malignancies. As immunohistochemistry (IHC) is cost-effective and broadly available, we applied the UWO3 score to a tissue microarray (TMA) consisting of 197 independent HPV^+^ HNSCC patients (**Supplementary Figure S4A**). Immune desert patients experienced inferior DFS (HR = 3.1, 95% CI 1.1 – 9.7, *P* = 0.038, univariate Cox model; **Figure 1A, Supplementary Figure S4B**) compared to the immune rich patients, highlighting the potential of the UWO3 immune classification as to be implemented as an IHC assay.

Finally, to validate the association between UWO3 score and survival outcomes, we performed blinded validations in a retrospective cohort and a prospective cohort. Patients were assigned to immune classes using UWO3 before unmasking of clinical outcomes by our external collaborators. In a retrospective cohort of HPV^+^ HNSCC patients (n = 262) treated primarily with surgery at Washington University at St. Louis (WashU) and Vanderbilt University^31^, the immune classes exhibited distinct DFS (*P* = 0.01, log rank test; immune desert *vs*. immune rich: HR = 2.5, 95% CI 1.3 – 4.7, *P* = 0.004, univariate Cox model; **Figure 1A,C, Supplementary Figure S5A**) and OS (*P* = 0.01, log rank test; immune desert *vs*. immune rich: HR = 3.44, 95% CI 1.59 – 7.44, *P* = 0.002, univariate Cox model; **Figure 1B,D, Supplementary Figure S5A**). In the prospective Big Data and Models for Personalized Head and Neck Cancer Decision Support (BD2Decide) study (n = 286, NCT02832102) of 286 locoregionally-advanced p16-positive patients treated homogeneously with radiation and/or chemotherapy at seven European institutions^32^, the immune classes exhibited distinct DFS (*P* = 0.03, log rank test; immune desert *vs*. immune rich: HR = 2.6, 95% CI 1.1 – 5.7, *P* = 0.02, univariate Cox model; **Figure 1A,C, Supplementary Figure S5C**) and OS (*P* = 0.004, log rank test; immune desert *vs*. immune rich: HR = 5.0, 95% CI 1.6 – 15, *P* = 0.004, univariate Cox model; **Figure 1B,D, Supplementary Figure S5D**). Taken together, we have shown that the UWO3 immune class is robustly associated with survival outcomes in six independent cohorts across different profiling platforms and geographic jurisdictions.

### Pooled analysis of all cohorts underscores UWO3 immune class as a strong independent predictor

As the association between immune groups and survival outcomes for each cohort were homogeneous between cohorts (*I*^2^ < 50%, *P* >0.10; **Figure 1A-D**), we performed a pooled analysis of all six cohorts (LHSC, TCGA, JHU, TMA, WashU, and Vanderbilt, and BD2Decide; total n = 906). The immune class defined by UWO3 was strongly associated with DFS (*P* = 4 × 10^−8^, log rank test, **Figure 1E**) and OS (*P* = 9 × 10^−7^, log rank test, **Figure 1F**) in this pooled cohort.

The 5-year DFS probabilities for the immune rich, mixed, and immune deserts group were 88.1%, 75.7%, and 67.3%, respectively. Immune desert patients exhibited inferior DFS over the immune rich (HR = 3.23, 95% CI 2.1 – 4.9, *P* = 2 × 10^−8^, univariate Cox model; **Figure 1E**) and mixed (HR = 1.38, 95% CI 1.0 – 1.9, *P* = 0.041, univariate Cox model; **Figure 1E**) patients. The mixed patients also exhibited worse DFS over the immune rich patients (HR = 2.34, 95% CI 1.5 – 3.6, *P* = 6 × 10^−5^, univariate Cox model; **Figure 1E**). The 5-year OS probabilities for the immune rich, mixed, and immune deserts group were 90.4%, 79.3%, and 71.8%, respectively. Immune desert patients exhibited inferior OS over the immune rich (HR = 2.89, 95% CI 1.9 – 4.4, *P* = 7 × 10^−7^, univariate Cox model; **Figure 1F**) and mixed (HR = 1.40, 95% CI 1.0 – 1.9, *P* = 0.041, **Figure 1F**) patients. The patients with mixed tumours also exhibited worse OS than immune rich patients (HR = 2.05, 95% CI 1.3 – 3.2, *P* = 0.0011, univariate Cox model; **Figure 1F**). The survival differences by UWO3 immune group persisted for patients undergoing both primary surgery (n=324. Immune rich *vs*. immune desert DFS: HR = 3.12, 95% CI 1.7 – 5.9, *P* = 0.0004, univariate Cox model; **Supplementary Figure S6 A, B**) or primary radiation (n=293. Immune rich *vs*. immune desert DFS: HR = 4.81, 95% CI 2.0 – 11.4, *P* = 0.0003, univariate Cox model; **Supplementary Figure S6 C, D**).

In a multivariate Cox proportional hazards model stratified for cohort, the association between the UWO3 immune class and DFS was independent of other clinical factors (Immune desert *vs*. immune rich: HR = 9.0, 95% CI 3.17 – 25.5, *P =* 3.6 × 10^−5^; Mixed *vs*. immune rich: HR = 6.4 (95% CI 2.2 – 18.7, *P =* 0.0006; **Table 1**). Brier score analysis demonstrates that the UWO3 immune group alone had a lower prediction error for disease-free survival than a Cox model of clinical factors (AJCC8 stage, sex, smoking status, and age) (*P* = 0.049, *χ*^2^ test; **Figure 2A**). Furthermore, integration of the UWO3 immune group with other clinical factors (UWO3 + Full Clinical) further decreased prediction error (*P* = 5 × 10^−7^, *χ*^2^ test; **Figure 2A**). We analyzed the relative contribution of each parameter to predict DFS and identified UWO3 immune group (50.9%, **Figure 2B**) as the strongest parameter, compared to other clinical factors. Thus, UWO3 immune class is a strong independent prognostic factor that can improve the risk-stratification of HPV^+^ HNSCC.

**Table 1.**
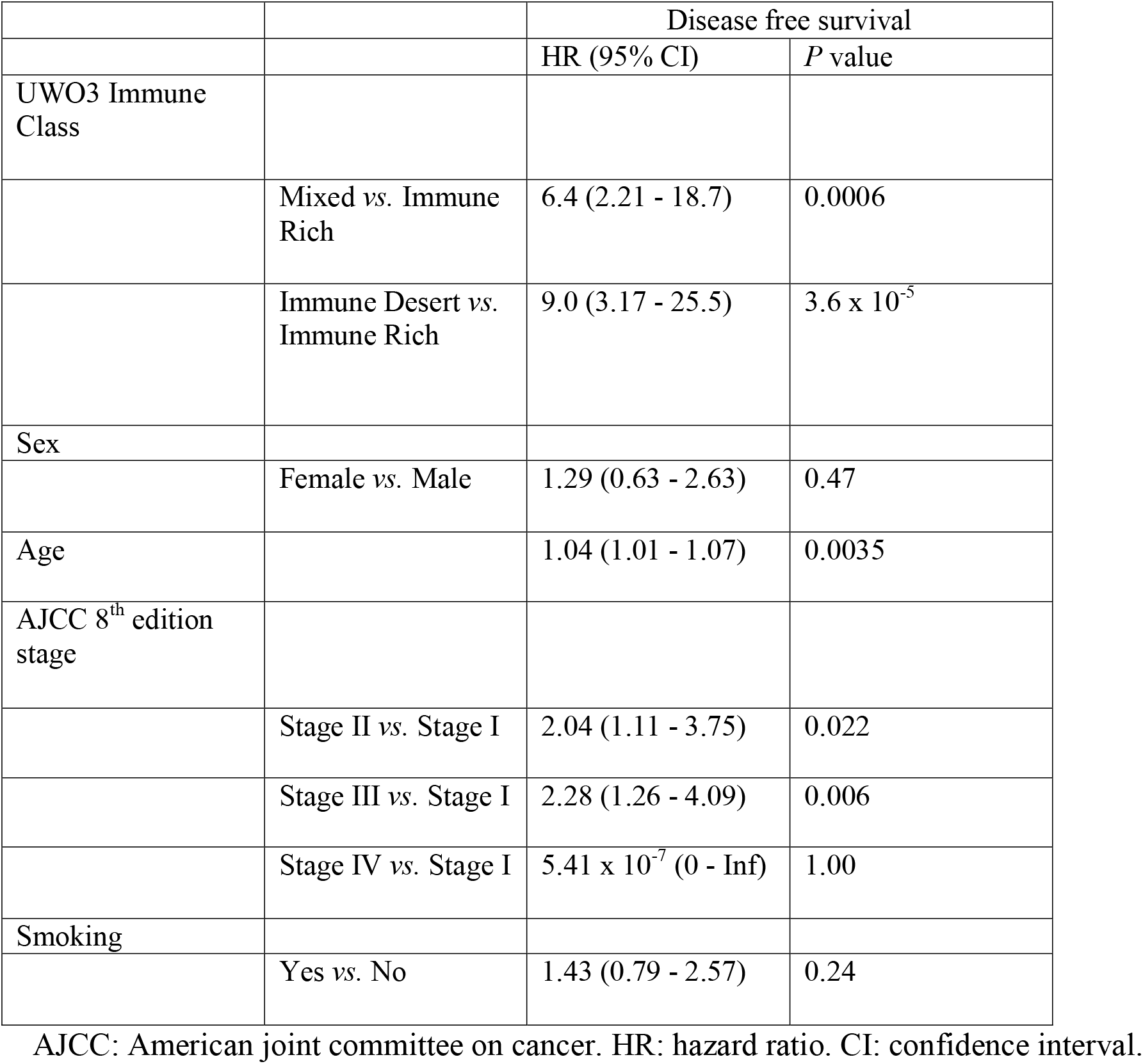
Multivariate analysis of the UWO3 immune class.

|  |  | Disease free survival |  |
| --- | --- | --- | --- |
|  |  | HR (95% CI) | P value |
| UWO3 Immune Class |  |  |  |
|  | Mixed vs. Immune Rich | 6.4 (2.21 - 18.7) | 0.0006 |
| | Immune Desert vs. Immune Rich | 9.0 (3.17 - 25.5) | $3.6 \times 10^{-5}$ |
| Sex |  |  |  |
|  | Female vs. Male | 1.29 (0.63 - 2.63) | 0.47 |
| Age |  | 1.04 (1.01 - 1.07) | 0.0035 |
| AJCC 8<sup>th</sup> edition stage |  |  |  |
|  | Stage II vs. Stage I | 2.04 (1.11 - 3.75) | 0.022 |
|  | Stage III vs. Stage I | 2.28 (1.26 - 4.09) | 0.006 |
| | Stage IV vs. Stage I | $5.41 \times 10^{-7}$ (0 - Inf) | 1.00 |
| Smoking |  |  |  |
|  | Yes vs. No | 1.43 (0.79 - 2.57) | 0.24 |

AJCC: American joint committee on cancer. HR: hazard ratio. CI: confidence interval.

**Figure 2.**
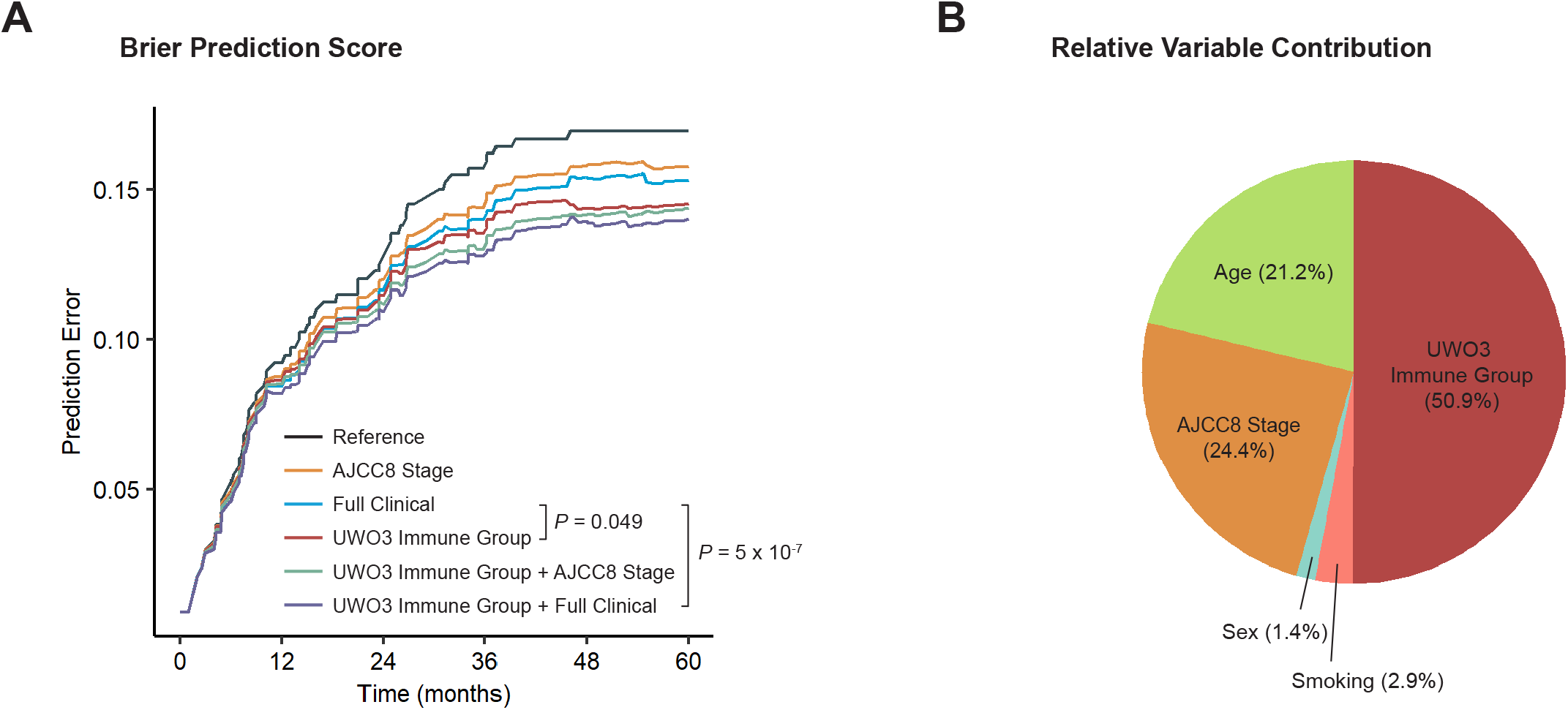
UWO3 immune class outperforms clinical factors in predicting disease-free survival. Brier prediction score analysis (**A**) shows lower error rate, thus higher prediction accuracy of disease-free survival for the UWO3 immune class than major clinical factors combined (AJCC8 stage, age, sex, smoking status). Integration of UWO3 immune group with other clinical factors further decreased prediction error rate. Relative importance of each risk (**B**) parameter to survival risk using the Pearson *χ*^2^ test for clinical parameters plus UWO immune group shows that immune group is the most important factor. AJCC: American Joint Committee on Cancer.

### UWO3 immune classification has implications for immunotherapy and treatment de-intensification

As the immune rich patients exhibit excellent survival outcomes, we hypothesized that the UWO3 score can identify patients who respond favourably to de-intensified treatment. We used RNA-seq data from the phase II MC1273 (NCT01932697) trial^17^, which tested an aggressive de-escalation regimen of 30Gy radiation with concurrent docetaxel post-surgery, and the 30ROC trial (NCT00606294)^33^, in which patients received 30Gy radiation and cisplatin. Higher UWO3 score was associated with higher odds of recurrence following aggressive treatment de-escalation (Odds ratio: 24.9, *P* = 0.0147, logistic regression with UWO3 as continuous variable stratified for cohort; **Figure 3**). Strikingly, 7 out of 9 patients (77.8%, **Figure 3**) in the immune desert group developed recurrence, while only 4 out of the 24 patients (16.6%) in the immune rich and mixed recurred. While limited by the sample size, these results support the potential ability of the UWO3 score to identify patients who will maximally benefit from aggressive treatment de-escalation.

**Figure 3.**
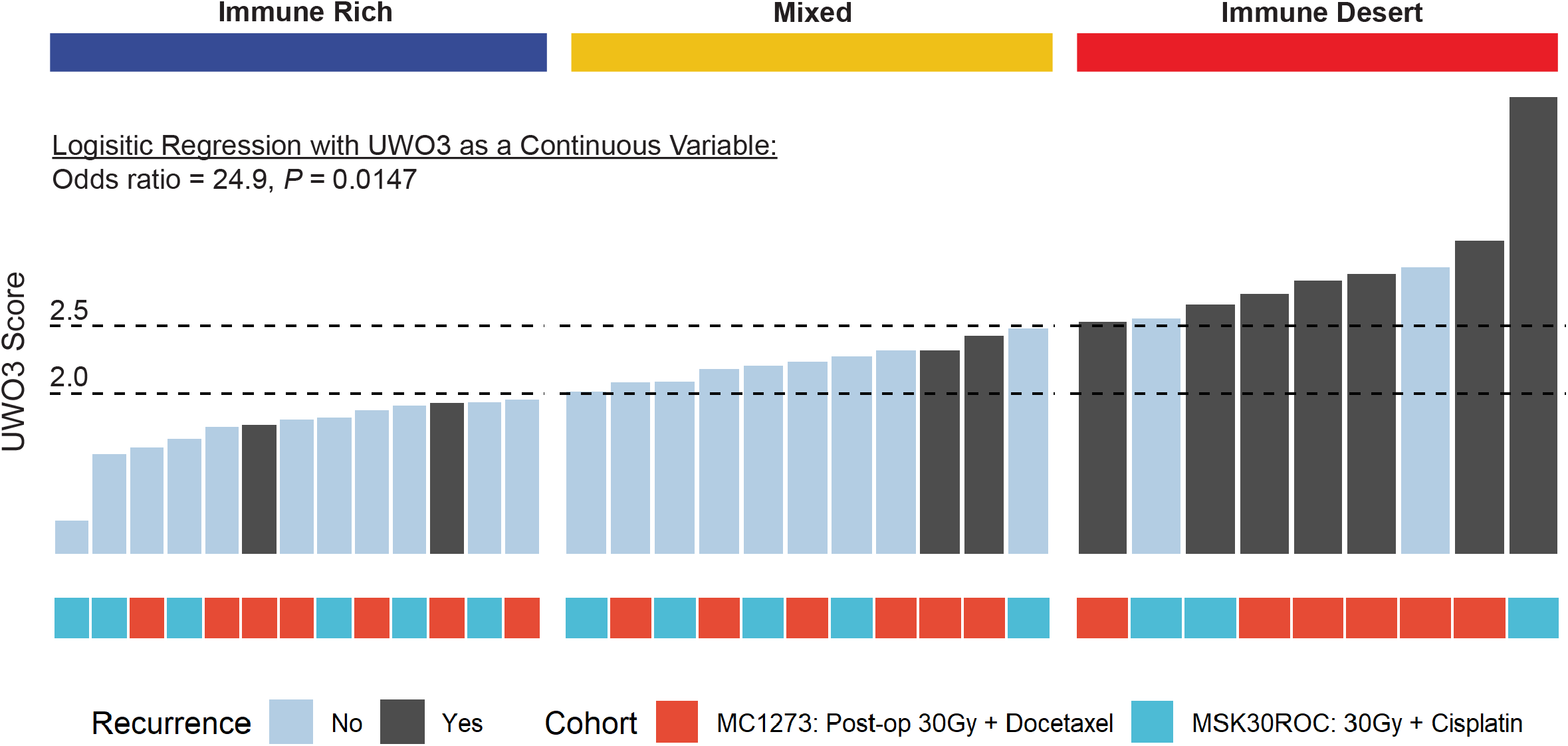
UWO3 immune classification has implications for immunotherapy and treatment de-intensification. UWO3 score can identify patients to aggressive radiation de-escalation from 70 grays (Gy) to 30 Gy in the Mayo Clinic MC1273 trial (NCT01932697) and the Memorial Sloan Kettering (MSK, NCT00606294) 30ROC trial. Recurrence is defined as patients who have not developed locoregional or metastatic disease as of last follow-up. Odds ratio and *P*-value are from logistic regression with UWO3 as a continuous variable and stratified for cohort.

## Discussion

Although HPV^+^ HNSCC patients have improved prognosis over their HPV-negative counterparts, a significant portion of patients still recur after initial treatment and are at risk of death. We demonstrate that the pre-treatment TME has dramatic effects in determining the prognosis of HPV^+^ HNSCC patients. Further, we describe a clinically translatable, extensively validated UWO3 immune classification tool that may allow biomarker-driven individualized treatment in HPV^+^ HNSCC. (**Figure 4**).

**Figure 4.**
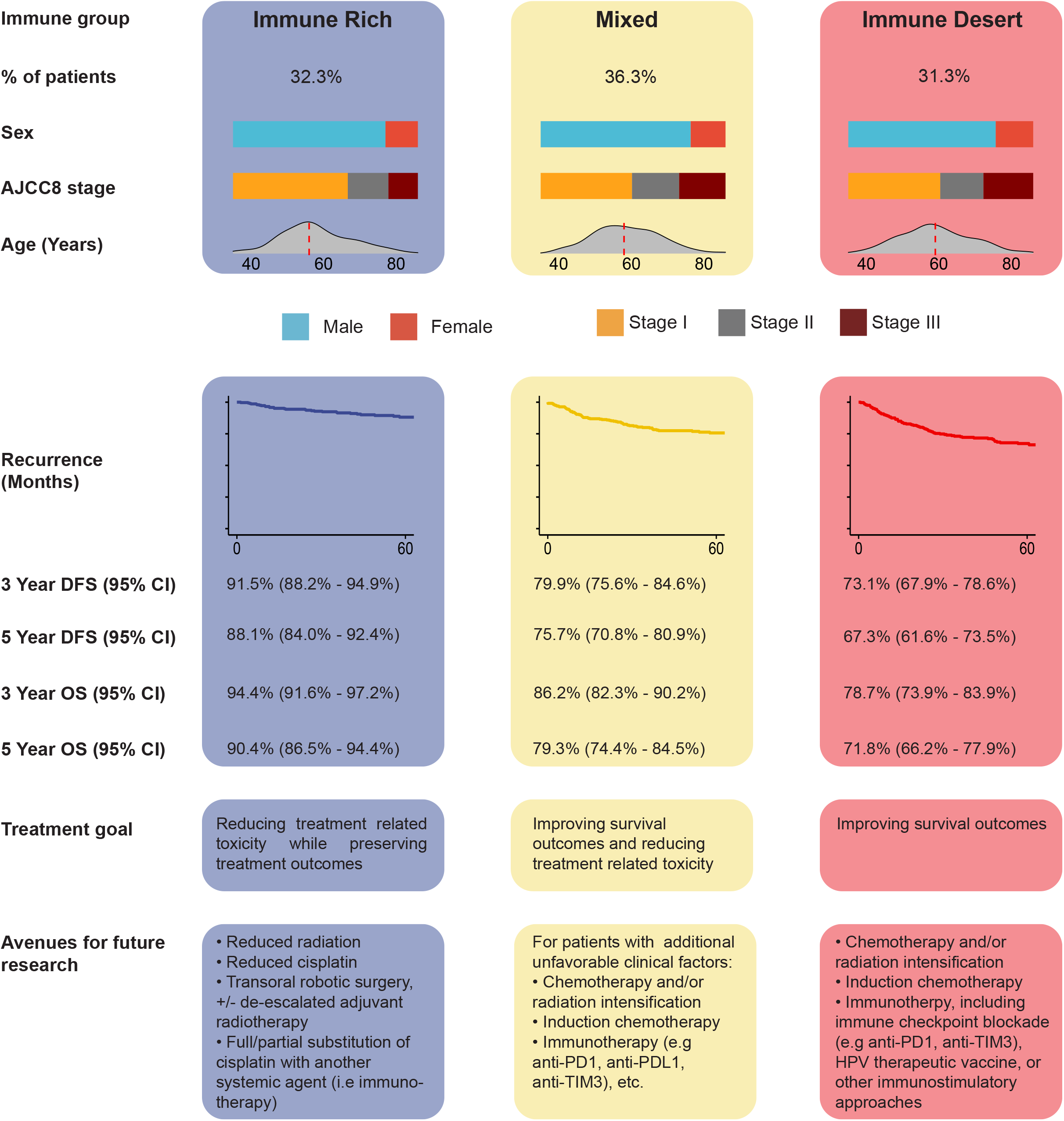
UWO3 immune classification of HPV^+^ HNSCC with implications for treatment de-intensification and immunotherapy. CI: confidence interval; DFS: disease-free survival; OS: overall survival.

The proposed immune classification reflects HPV^+^ HNSCC biology. CD3E is part of the T-cell receptor complex and its down-regulation on T-cells has been linked to worse prognosis in HNSCC^34^. ZAP70 plays important role in T-cell receptor signaling but is also highly expressed on NK-cells^35^. IRF4 directs the development, affinity maturation, and terminal differentiation of B cells, but also plays important roles in monocyte differentiation^36-40^. Accumulating evidence suggests that rich immune infiltration distinguishes HPV^+^ HNSCC from both HPV-negative HNSCC and cervical cancer, a cancer type with similar HPV-viral etiology^41^. There could be several reasons underlying the difference in the TME specifically in HPV^+^ HNSCC. The oropharynx region includes the Waldeyer’s tonsillar ring of lymphoid organs that are densely infiltrated with immune cells. Furthermore, HPV^+^ HNSCC is virally-driven and thus expresses unique viral antigens. The presence of viral-host fusion proteins following HPV integration in some tumours may be strongly immunogenic and stimulate anti-tumour immunity^42^. Recent work using single-cell RNA-sequencing has identified high proportions of HPV antigen-specific tumour infiltrating lymphocytes in HPV^+^ HNSCC^43,44^, including a subset of PD1^+^ TIM3^+^ (the protein encoded by the gene *HAVCR2*) terminally differentiated CD8 T-cells within HPV^+^ HNSCC tumour that fail to proliferate following antigen stimulation. Multiple studies are ongoing to assess the relevance of TIM3 inhibition in a variety of tumour types^45,46^. Further work will be needed to investigate whether the addition of immunotherapy (such as anti-PD1 and/or anti-TIM3) to CRT can stimulate anti-tumour immunity and improve survival outcomes for HPV^+^ HNSCC patients with an immune desert TME.

Patients with immune rich pre-treatment tumours have improved prognosis compared to the mixed and immune desert groups consistently across six different cohorts regardless of treatment. Thus, patients who are immune rich and have favorable clinical factors may be ideal candidates for aggressive treatment de-intensification. The inflammatory TME of the immune rich group and dense infiltration of PD1^+^ CD8 T cells pre-treatment supports further exploration of substitution of chemotherapy with immunotherapy in de-escalation settings. The three-arm phase III randomized controlled trial NRG-HN005 (NCT03952585) is currently evaluating such a regimen (60 Gy plus nivolumab) against the standard treatment of 70 Gy with cisplatin and a de-intensified regimen of 60 Gy plus cisplatin. Furthermore, the association between patients who have immune desert pre-treatment tumours and poor survival outcomes supports the investigation of neoadjuvant immunotherapy approaches, which have been found to increase T-cell density within HNSCC tumour^47^.

The key limitation of our study is that the treatment protocols delivered in each cohort were not uniform. However, the strong association of UWO3 with DFS in six cohorts, which spans different geographic jurisdictions, treatment methods, and patient populations suggests that the immune classes can treatment-agnostically predict survival outcomes. Another limitation is that our analysis of immune co-receptors abundance was performed using bulk RNA profiling and thus cannot distinguish the specific cell types expressing these immune checkpoint receptors. Moreover, some of these molecules (such as TIM3) are expressed on T-cells as well as dendritic cells and macrophages^48-50^. Nevertheless, recent work in HPV^+^ HNSCC using single-cell RNA-seq demonstrated that T-cells make up the majority of immune cell populations within the TME and are the major contributors of these molecules^24,51^. As single-cell technologies become more accessible, cohort-level studies to fully dissect HPV^+^ HNSCC and characteristics associated with survival will undoubtedly pave the way for a more nuanced understanding of HPV^+^ HNSCC TME.

The ideal patient population for treatment de-intensification and intensification in HPV^+^ HNSCC remains controversial^13^. Our retrospectively and prospectively-validated immune classification provides a readily implemented means to identify patients that may maximally benefit from treatment de-escalation or intensification. We are in the process of testing these hypotheses through biomarker-driven randomized controlled de-intensification and intensification trials.

## Methods

### Patient cohort

The study was approved by the Research Ethics Boards at Western University (REB 7182) and informed consent was obtained from each patient. Primary site fresh tumour samples were prospectively collected from patients with HPV^+^ oropharyngeal squamous cell cancer at Victoria Hospital, London Health Science Centre, London, Ontario, Canada between 2010 and 2016. Patient demographics and survival outcomes were prospectively collected. Frozen section analysis was carried out to confirm tumor cellularity greater than 70%. HPV status was confirmed via p16 immunohistochemistry as well as PCR and Sanger sequencing. Detailed clinical information in the LHSC cohort is provided in **Supplementary Table S1**. Disease free survival was defined as time from diagnosis to recurrence at any site or death. Recurrence was defined as the presence of local, regional, or distant disease after completion of treatment. Detailed descriptions of all other cohorts have been provided elsewhere^30-32,52,53^. Treatments received for each cohort are described in **Supplementary Table S2**. The reporting of the study followed guidelines from Reporting Recommendations for Tumor Marker Prognostic Studies (REMARK)^54^.

### RNA-seq processing, sequencing, and bioinformatics

Detailed information about RNA-seq sample preparation, processing, bioinformatic analyses, and the development and validation of the UWO3 score can be found in the **Extended Methods**. An online UWO3 calculator is available (https://www.nicholslab.com/uwo3calculator).

### Statistical analysis

All statistical analyses were performed with R software (v4.0.5) using the following packages: survival (v3.2.10), ggpubr (v0.4.0), stats (v4.0.5), and rms (v6.2-0). Wilcoxon rank sum test (2 categories) and Kruskal-Wallis test (>2 categories) were used to examine all relationships between categorical variables and quantitative variables. *P*-values were corrected for multiple testing using the Benjamini-Hochberg method. Fisher’s method was used to combine Wilcoxon rank sum test *P-*values across cohorts. The Fisher’s Exact test was used to analyze contingency tables. Survival analyses were analyzed with the log-rank test and Cox proportional hazards model with the survival (v3.1-12) package. The proportional hazards assumption was assessed using Schoenfeld residuals. Hazard ratio in the JHU cohort was estimated using Cox Regression with Firth’s penalized likelihood^55^, implemented using the coxphf package (v1.13.1), as monotone likelihood is observed due to no events in the immune rich group. Meta-analysis of the association between UWO3 immune class and disease-free survival using the inverse variance method through the R package meta (v5.1-1) using log_10_HR and SEM in a fixed-effect model. The significance of any discrepancies in the estimates of the treatment effects from the different cohorts was assessed using Cochrane’s test for heterogeneity and the *I*^2^ statistic as described previously^56^. Heterogeneity was considered statistically significant if the *P* value was less than 0.10 for the *χ*^2^ test. Brier’s error analysis of Cox models was calculated using the package pec (v2021.10.11). The relative importance of each parameter to survival risk was assessed using the *χ*^2^ from R package rms (v6.2-0). All tests were 2-sided.

## Data Availability

Raw data for the LHSC and TMA cohort available via request to ACN. Data from the TCGA HNSC cohort included in the current study is available from the Genomic Data Commons Data Portal (https://portal.gdc.cancer.gov/projects/TCGA-HNSC). Data from the JHU, Washington University at St. Louis, and BD2Decide cohort could be accessed from GEO with accession numbers GSE112026, GSE171898, and GSE163173. Data from the MC1273 and 30ROC trials could be accessed at GSE157517.

## Notes

### Prior presentations

None.

### Author contributions

P.Y.F.Z, M.J.C, M.S, M.I.K, J.W.B, A.C.N designed the study and experiments. P.Y.F.Z, M.J.C, M.S-T, L.D.C, X.L performed the bioinformatic analysis. J.W.B performed RNA extraction. M.S.S, S.C, L.L, F.H, R.H.B, C.R.L, K.S, T.P, X.W, F.L, E.P, C.P, S.D.DS, M.H, J.C.D, P.B., D.M, C.J.H, W.S, J.Y., K.F., D.A.P., P.L, S.D.S, A.M, P.L, E.W, H.Z, A.A, A.C.N provided patients materials and clinical data. P.Y.F.Z, M.S, M.I.K, J.W.B, S.S-C.L,A.A, P.C.B, J.S.M, and A.C.N discussed data and wrote the text. P.C.B, J.S.M, and A.C.N supervised the study. All authors approved the manuscript before submission.

## Acknowledgements

ACN was supported by the Wolfe Surgical Research Professorship in the Biology of Head and Neck Cancers Fund. PYFZ is supported by a CIHR Vanier Canada Graduate Scholarship and a PSI Foundation Fellowship.

---

**Supplementary Figure S1.**
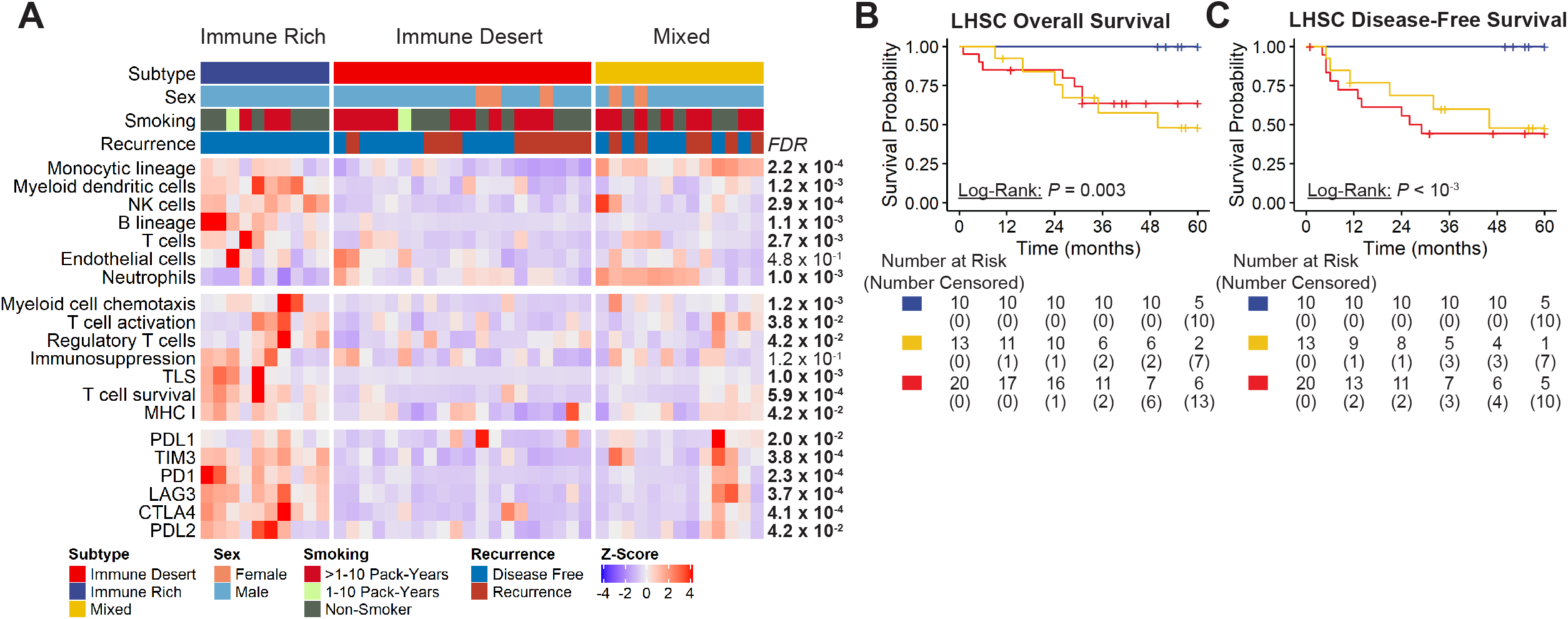
Immune classification of HPV^+^ HNSCC. Composition of the HPV^+^ HNSCC TME (**A**) as defined by MCP-counter generated Z-score, immune gene signature scores, and expression immune checkpoint related genes. Clustering based on K-means clustering of MCP-counter estimated immune abundance Z-score. *Q* values derived from Benjamini-Hochberg corrected Kruskal-Wallis *P* values. Kaplan-Meier analyses of overall survival (**B**) and disease free survival (**C**) of HPV+ oropharyngeal cancer patients by their tumor immune group show distinct survival patterns. All tests are two-sided log-rank tests.

**Supplementary Figure S2.**
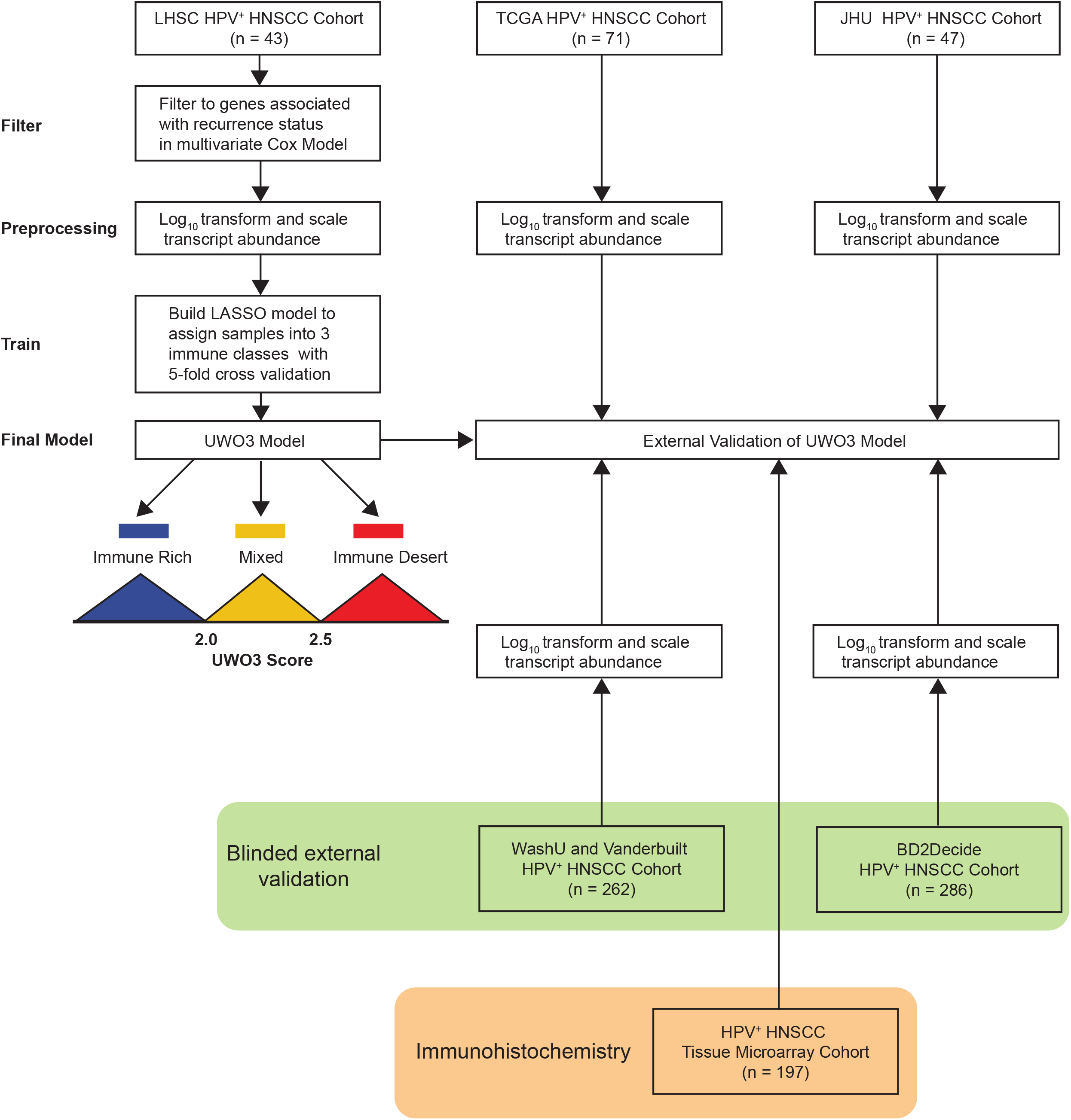
Derivation and validation of the UWO3 score.

**Supplementary Figure S3.**
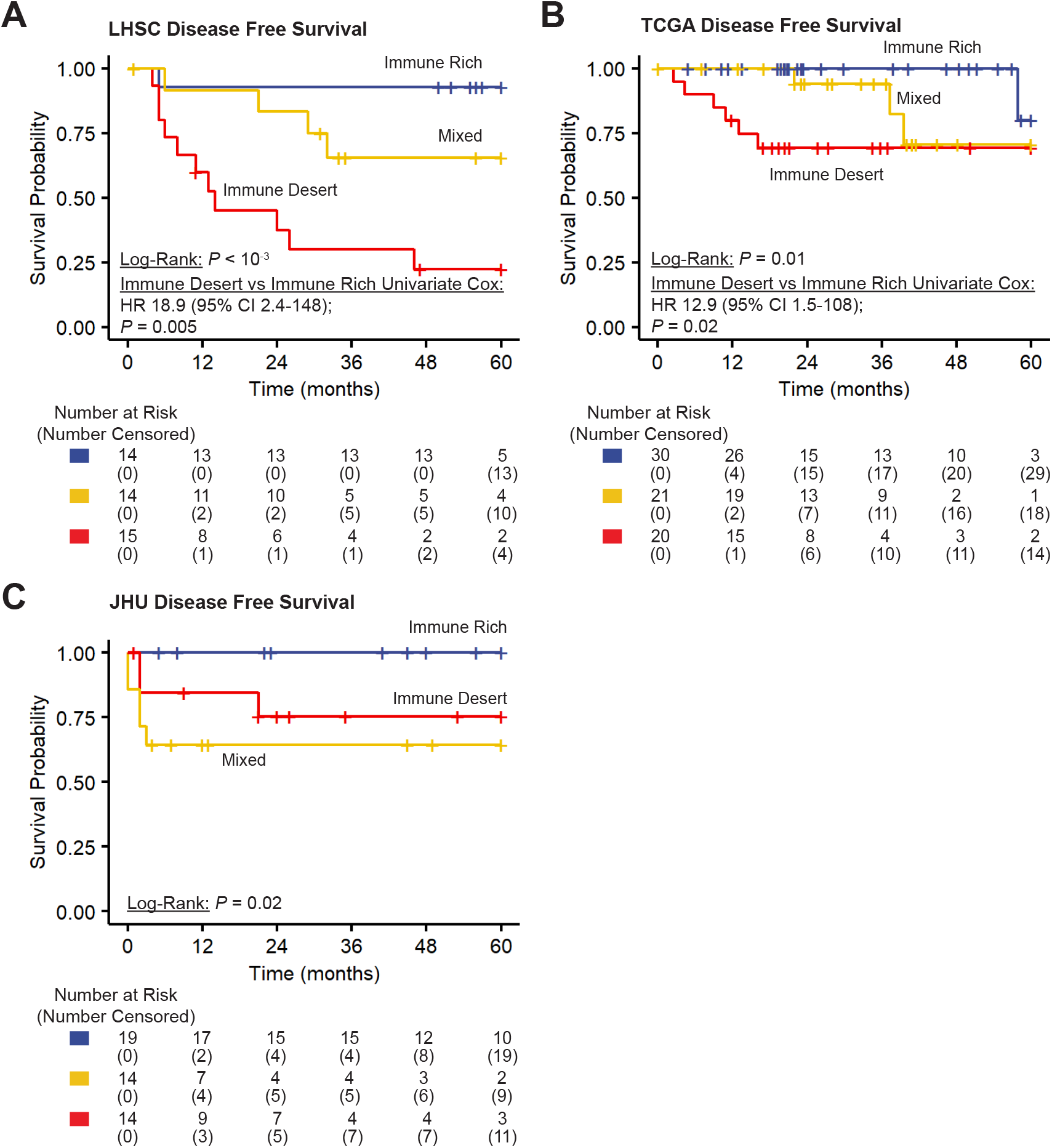
UWO3 immune groups predict treatment response in two external cohorts. Kaplan-Meier analyses of disease-free survival of HPV+ HNSCC patients in the LHSC (**A**), TCGA (**B**), and JHU (**C**) cohort by their immune group as assigned through the UWO3 score demonstrate that UWO3 immune groups can predict survival. *P* values from two-sided log rank tests and Cox proportional regression model.

**Supplementary Figure S4.**
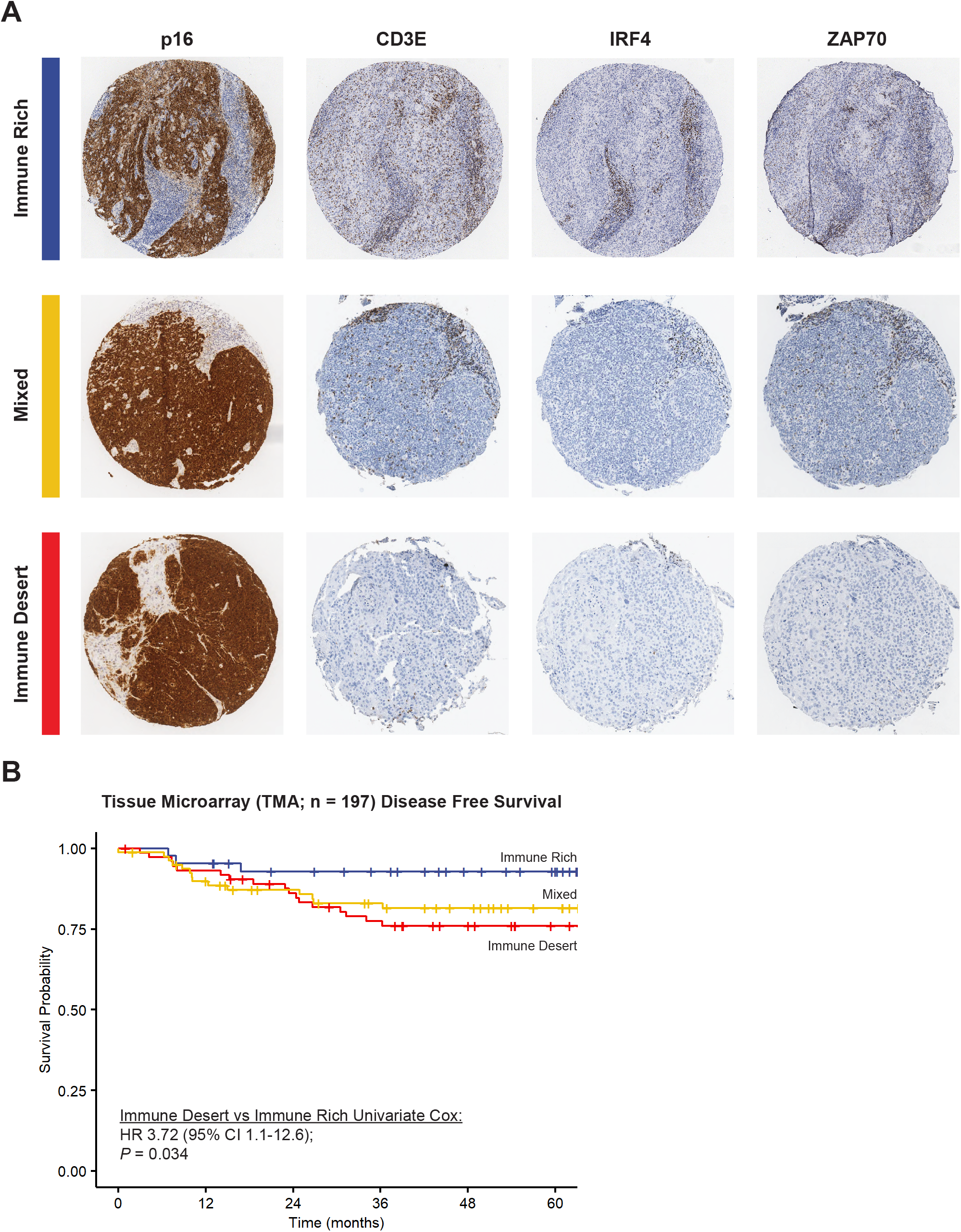
UWO3 immune groups predict recurrence in a cohort of HPV^+^ HNSCC patients using immunohistochemistry. Representative TMA images stained for hematoxylin and eosin (H&E), p16 (a surrogate marker of HPV status), and the three proteins used to calculate UWO3 (CD3E, ZAP70, and IRF4, **A**). Kaplan-Meier analyses of the disease-free survival (**B**) of the tumour microarray (TMA) demonstrate that UWO3 immune groups defined using clinically-validated antibodies can predict recurrence in HPV^+^ HNSCC. *P* value from Cox proportional regression model.

**Supplementary Figure S5.**
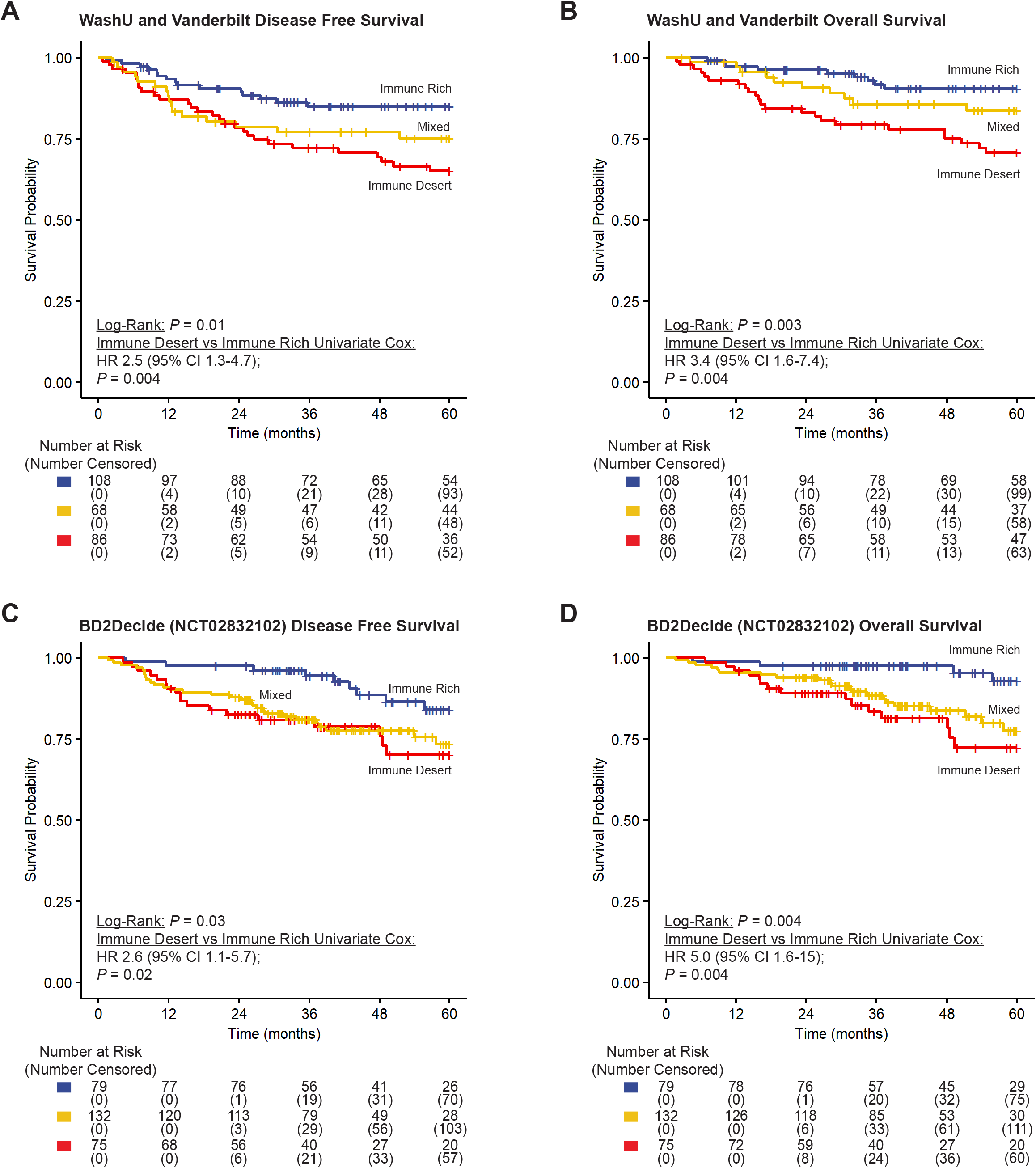
Blinded external validation of the UWO3 immune groups in a retrospective cohort and a prospective cohort. Kaplan-Meier analyses of disease-free survival (**A**) and overall survival (**B**) of HPV+ HNSCC patients in a retrospective cohort of patients from the Washington University at St. Louis (WashU) and Vanderbilt University shows that UWO3 immune groups are associated with distinct survival outcomes. Further, in a prospective cohort (BD2Decide, NCT02832102) where HPV+ HNSCC patients received uniform treatment, Kaplan-Meier analyses of disease-free survival (**C**) and overall survival (**D**) shows that UWO3 immune groups are associated with distinct survival outcomes. *P* values from two-sided log rank tests and Cox proportional regression model.

**Supplementary Figure S6.**
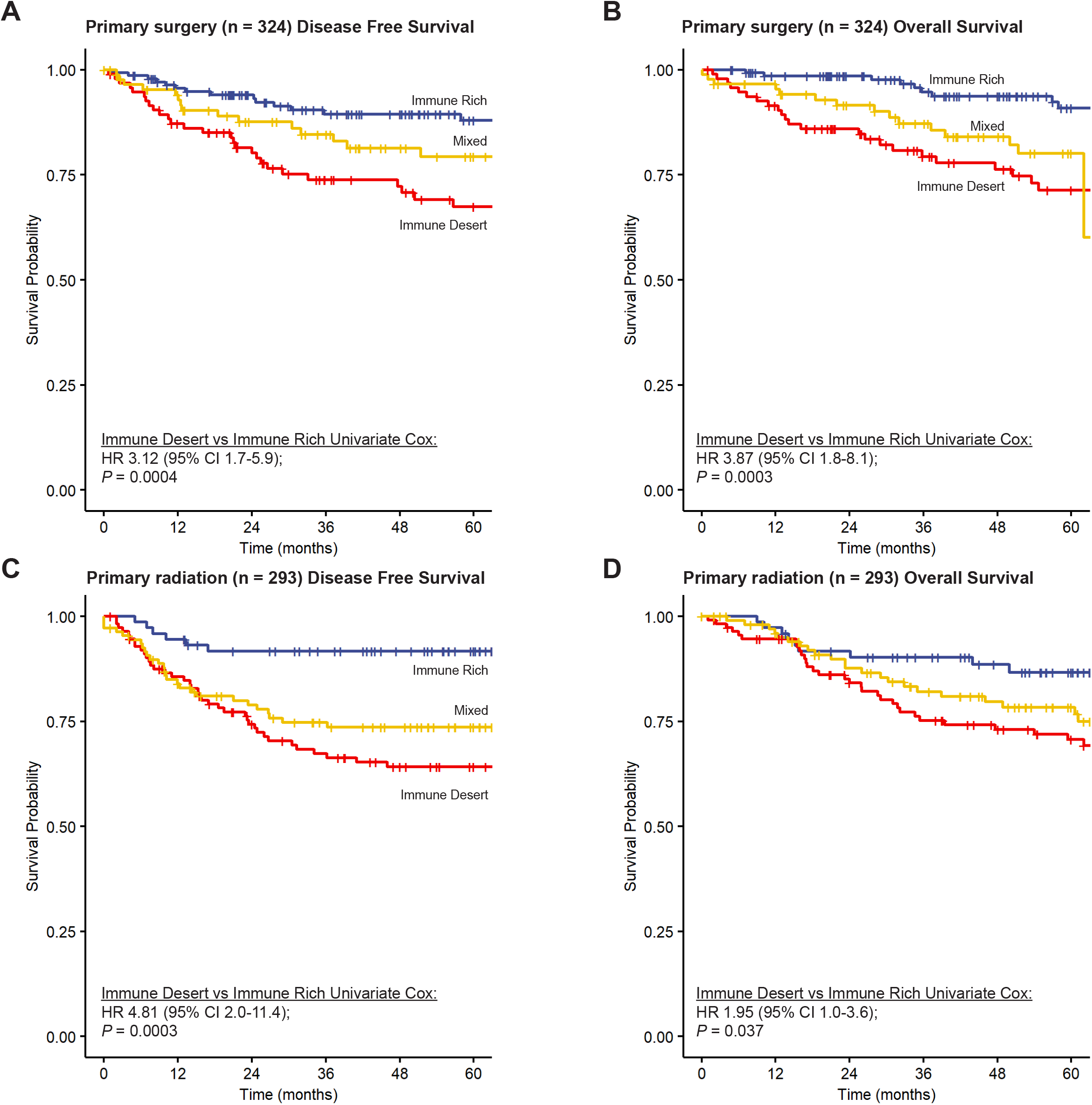
UWO3 score can predict survival outcomes independent of primary treatment. Kaplan-Meier analyses of disease-free survival (**A**) and overall survival (**B**) of HPV+ HNSCC patients treated with primary surgical approach with or without adjuvant therapy. Kaplan-Meier analyses of disease-free survival (**C**) and overall survival (**D**) of HPV+ HNSCC patients treated with primary radiation with or without chemotherapy. *P* values from univariate Cox proportional regression model.

