## Extended Methods for "A Clinically Translatable Immune-based Classification of HPV-associated Head and Neck Cancer with Implications for Biomarker-Driven Treatment Deintensification and Immunotherapy"

**Supplementary Methods**

**RNA Processing and Sequencing**

Total RNA and DNA was isolated using Qiagen AllPrep DNA/RNA kits. HPV status was confirmed by real time PCR as we have previously described^1,2^. One microgram of total RNA was shipped to The Center for Applied Genomics (Hospital for Sick Children, Toronto, ON) for quality control, library preparation, and sequencing. RNA quality was confirmed with a Bioanalyzer and libraries were prepared using a NEB Ultra II Directional mRNA library kit. Samples were then processed using random primers and sequenced using an Illumina HiSeq 2500 paired end for 50-90 million reads/sample (median: 66 million reads, **Supplementary table S1**).

FASTQ files were pre-processed with trim_galore (v0.6.4) and then quality controlled using FastQC (v0.11.9). Each sample were mapped to the human reference genome GRCh38 (v97) using STAR aligner (v2.7.2b) in two-pass mode^3^, and quantified using HTSeq-count (v0.12.3) intersection-strict mode^4^. Read normalization and differentially expressed gene testing was conducted using DESeq2 (v1.26.0)^5^. Differentially abundant transcripts between the disease-free and recurrent patient are defined as transcripts having at least an average of normalized reads of 10, an absolute log_2_ fold change (log_2_FC) greater than 2, and a Benjamini-Hochberg adjusted p-value of less than 0.05. Generation and processing of external cohorts have been described elsewhere^6-10^.

**HPV genotyping *via* HPV transcript quantification**

HPV genotyping were performed on raw RNA-seq reads using HPViewer (branch c62f29e, available at https://github.com/yuhanH/HPViewer), on a database of 182 repeat masked HPV strains. HPV reads were then quantified using HTSeq-count intersection-strict mode to the subtype with the highest read-number^11^.

**TME Estimation**

The TME composition of each sample was estimated using the MCP-counter score (v1.1.0)^12^. The score was based on previously analyzed transcriptomic markers that are found to be characteristic of the specific immune population and were proportional to the abundance of each cell population within the tumour. Comparison with other immune deconvolution methods have found the method to be highly accurate and capable of inter-sample comparisons^13^. The MCP-counter signatures composition are as follows: T cells: CD28, CD3D, CD3G, CD5, CD6, CHRM3-AS2, CTLA4, FLT3LG, ICOS, MAL, PBX4, SIRPG, THEMIS, TNFRSF25 and TRAT1; B lineage: BANK1, CD19, CD22, CD79A, CR2, FCRL2, IGKC, MS4A1 and PAX5; natural killer cells: CD160, KIR2DL1, KIR2DL3, KIR2DL4, KIR3DL1, KIR3DS1, NCR1, PTGDR and SH2D1B; monocytic lineage: ADAP2, CSF1R, FPR3, KYNU, PLA2G7, RASSF4 and TFEC; myeloid dendritic cells: CD1A, CD1B, CD1E, CLEC10A, CLIC2 and WFDC21P; neutrophils: CA4, CEACAM3, CXCR1, CXCR2, CYP4F3, FCGR3B, HAL, KCNJ15, MEGF9, SLC25A37, STEAP4, TECPR2, TLE3, TNFRSF10C and VNN3; endothelial cells: ACVRL1, APLN, BCL6B, BMP6, BMX, CDH5, CLEC14A, CXorf36 (also known as DIPK2B), EDN1, ELTD1, EMCN, ESAM, ESM1, FAM124B, HECW2, HHIP, KDR, MMRN1, MMRN2, MYCT1, PALMD, PEAR1, PGF, PLXNA2, PTPRB, ROBO4, SDPR, SHANK3, SHE, TEK, TIE1, VEPH1 and VWF.

**Clustering**

K-means clustering was performed on Z-scores of the immune cell abundance estimation (**Supplementary Figure 1A**) using the kmeans wrapper of ComplexHeatmap package (v2.1.0) when generating heatmap^14^. Kmeans was run 1000 times to generate consensus k-means clustering. The number of K clusters was selected using the silhouette method through the fviz_nbclust function of the factoextra R package (v1.0.6).

**Immune gene signatures**

Immune gene signatures were derived from other studies^15^. Briefly, each signatures were computed as geometric mean of the abundance of its included genes: immunosuppression (TGFB1, TGFB3, LGALS1, and CXCL12), regulatory T cells (FOXP3, TNFRSF18), T cell survival score (CD70 and CD27), T cell activation (CXCL9, CXCL10,CXCL16,IL15, and IFNG), myeloid chemotaxis(CCL2), MHC I (HLA-A, HLA-B, HLA-C, HLA-E, HLA-F, HLA-G, and B2M), and tertiary lymphoid structures (CXCL13).

**UWO3 score development and prediction**

Statistically significant transcripts between disease-free and recurrent tumour were filtered to only include transcripts whose abundance is independently associated with prognosis. The cohort was dichotomized into high and low abundance groups for each transcript and tested for association with prognosis in a Cox proportional hazard multivariate model including important clinical variables (age at diagnosis, sex, T stage, N stage, smoking status, alcohol abuse status). Transcripts with FDR < 0.25 had it abundance added 0.1, log-10 transformed, and scaled for score development. To facilitate the development of protein diagnostic tools for pathology, the gene lists were further filtered to contain genes for which the RNA abundance is highly correlated with its protein abundance (FDR < 0.05 & rho > 0.6, Spearman Correlation) using the HPV-negative HNSCC CPTAC cohortP^16^P.

For the development of UWO3 score to differentiate between the 3 TME signatures, we used a regularized linear regression technique based on the LASSO algorithm as implemented in the glmnet (v3.0-2) R package using 5-fold cross-validation. The score was trained with “gaussian” family of model to minimize mean squared error (MSE). A minimal subset of 3 genes (CD3E, IRF4, and ZAP70) was selected whose weighted RNA abundance (UWO3 score) was highly associated with survival outcomes in the training cohort. RNA-abundance for each of the gene is then log_10_, scaled within the cohort, and used to calculate the UWO3 score with the following equation: UWO3 = 2.23255813953488 + ZAP70 * -0.224179275535717 + IRF4 * -0.137196384259042 + CD3E * -0.273419927248369. Using UWO3, tumours are then assigned to an immune class as follow: UWO3 <= 2 were predicted be “immune rich”, 2 < UWO3 <= 2.5 were predicted to be “mixed”, and UWO3 > 2.5 were predicted to be “immune desert”. Cut offs were determined by maximizing the correct number of tumours assigned to the same immune subtype through both clustering and the UWO3 and ensuring similar number of patients between each group.

**Tumour microarray (TMA) and immunohistochemistry**

All samples were obtained with informed consent after approval of the Institutional Review Board at Western University, the University of Calgary, and the University of British Columbia. The TMA from University of Calgary and University of British Columbia have been described previously^9,17^. The TMA from Western University are processed as following. The formalin-fixed paraffin-embedded (FFPE) blocks for each tumor was sectioned and stained with hematoxylin & eosin (H&E) to confirm the presence of human tumor. A Manual Tissue Arrayer (MTA-1; Beecher Instruments Inc.) was used to punch out 3–4 cylindrical cores of 0.6 mm diameter from each tumour sample. Cores were arrayed into recipient paraffin blocks. Control tissues were also included on each block. Cores were sealed into recipient blocks by heating at 40 °C for ~40mins. Blocks were sectioned into 1.5 μM sections and affixed to glass slides. Every ninth slide was stained with H&E to provide a reference. Additional details are available in the MTA-1 Instruction Manual (www.beecherinstruments.com). IHC staining was completed at the Department of Pathology & Laboratory Medicine and the Molecular Pathology Core Facility (Western University). Tissues were examined using an Aperio ScanScope® slide scanner and staining quantification was performed using the QuPath (v0.2.3).

To translate the UWO3 score for use in TMAs, we stained the TMA with anti-CD3 (IR503, Agilent Dako), anti-ZAP70 (clone 2F3.2, IR653, Agilent Dako), and anti-IRF4 (clone MUM1p, IR644, Agilent Dako) antibodies on an Omnis staining platform (Agilent Dako). The tumor was contoured by a subspecialist pathologist and the number of cells within the tumour positive for each marker was quantified using the positive cell detection function in QuPath^18^. The percentage of positive cells within each tumour was then used to create Z-score and used to generate the UWO3 score. Using UWO3, tumours are then assigned to an immune class as follow: UWO3 <= 2 were predicted be “immune rich”, 2 < UWO3 <= 2.5 were predicted to be “mixed”, and UWO3 > 2.5 were predicted to be “immune desert”.

**Funding:**

This study was funded by Canadian Institute for Health Research grants MOP340674 to ACN and PCB, PJT-173496 to JSM and ACN, NIH/NCI award number P30CA016042 to PCB, European Union Horizon 2020 Framework Program Award Number: 689715, and AIRC ID 23573 t to LDC. ACN was supported by the Wolfe Surgical Research Professorship in the Biology of Head and Neck Cancers Fund. ALA was supported in part by a NIH/NIDCR Developmental Research Program (DRP) Grant from the Yale Head and Neck SPORE P50-DE030707.

**Data Availability:**

Raw data for the LHSC and TMA cohort available via request to ACN. Data from the TCGA HNSC cohort included in the current study is available from the Genomic Data Commons Data Portal (https://portal.gdc.cancer.gov/projects/TCGA-HNSC). Data from the JHU, Washington University at St. Louis, and BD2Decide cohort could be accessed from GEO with accession numbers GSE112026, GSE171898, and GSE163173. Data from the MC1273 and 30ROC trials could be accessed at GSE157517.
